# Next-Gen Intestinal Parasite Detection: Leveraging Metataxonomics for Improved Diagnosis of Intestinal Protists and Helminths

**DOI:** 10.1101/2025.08.03.25332887

**Authors:** Katherine Bedoya-Urrego, Nicolas Rozo-Montoya, Ana L. Galván-Díaz, Gisela M. Garcia-Montoya, Juan F. Alzate

## Abstract

Intestinal parasites continue to pose a significant public health burden in low- and middle-income countries and are increasingly recognized in developed regions. Traditional diagnostic methods, primarily based on microscopy, remain widely used despite limitations in sensitivity and taxonomic resolution. This study employs a next-generation sequencing (NGS)-based metataxonomic approach, integrated with classical phylogenetic methods, to characterize intestinal parasites in rural Colombian populations. We compare its performance with conventional microscopy, focusing on both protist and geohelminth detection. Our findings demonstrate that metataxonomics outperforms microscopy in detecting *Strongyloides stercoralis*, while *Trichuris trichiura* was more effectively identified via microscopy—likely due to resistant eggshells impeding DNA extraction. The metataxonomic approach also enabled confident species- and subtype-level classification, particularly for *Blastocystis* and *Entamoeba* spp., revealing high colonization rates and frequent mixed infections. This study underscores the diagnostic power of NGS-based metataxonomics, which proved highly effective for broad-spectrum parasite detection, while also highlighting areas for methodological improvement—especially in primer design to enhance taxonomic coverage.

## INTRODUCTION

Intestinal protist and helminth infections remain a significant public health challenge globally, particularly in developing countries, where poor sanitation, and limited access to safe drinking water, inadequate diagnostic capacity, and widespread socioeconomic vulnerability contribute to the transmission (Echazú et al., 2015; Getie et al., 2024; Strunz et al., 2014). These conditions result in persistent fecal contamination of soil, water, and food, facilitating the continued spread of these infections (González-Ramírez et al., 2022). Although governmental initiatives, often supported by the World Health Organization (WHO), have been implemented to curb transmission (World Health Organization, 2015, 2021), actions focused on health promotion and infection prevention and control remain insufficient and largely ineffective.

Globally, approximately 3.5 billion people are infected with intestinal protozoa and helminths, of whom around 450 million are symptomatic, leading to significant morbidity and mortality (Gadisa & Jote, 2019; Tegen et al., 2020). These parasites contribute substantially to gastrointestinal disorders such as diarrhea, chronic malabsorption, malnutrition, and impaired growth and development (Ahmed, 2023; Fauziah et al., 2022). The associated health consequences are particularly severe among vulnerable populations, including school-age children, pregnant women, and immunocompromised individuals. The resulting burden highlights the need for improved detection and control strategies.

Among intestinal protozoan parasites, *Giardia intestinalis*, *Entamoeba histolytica*, *Cryptosporidium* spp. and *Blastocystis* spp. are the most reported worldwide (Ahmed, 2023). In parallel, soil-transmitted helminths (STH) affect more than 1.5 billion people globally, accounting for nearly 24% of the world’s population (Krolewiecki et al., 2013). The most frequently reported species include *Ascaris lumbricoides*, *Trichuris trichiura*, and the hookworm *Ancylostoma duodenale* and *Necator americanus* (Mutombo et al., 2019; Savioli et al., 2017). Additionally, the threadworm *Strongyloides stercoralis*, while significant and likely underreported, is often considered separately (Mutombo et al., 2019). Zoonotic STH species such as *Ancylostoma ceylanicum*, *Ascaris suum*, *Trichuris vulpis*, and *Trichuris suis* have also increasingly been detected in humans, expanding the spectrum of helminths known to cause disease in the human host (Hassan et al., 2021; Khurana et al., 2021; Rodpai et al., 2018).

Microscopy-based methods are the common standard for intestinal parasite detection due to their low cost and simplicity (Momčilović et al., 2019). However, they exhibit low sensitivity and specificity which is highly dependent on the parasite load and technician expertise (Miswan et al., 2022). They also lack the resolution needed to accurately differentiate morphologically similar species, such as *Entamoeba histolytica* from non-pathogenic *Entamoeba* spp. (Carrero et al., 2020), *Ascaris lumbricoides* from closely related zoonotic species like *Ascaris suum* (Betson et al., 2014), or among human-infecting hookworms such as *Necator americanus* and *Ancylostoma duodenale* (Wang et al., 2012). Consequently, there is a growing need for studies using advanced diagnostic tools to accurately identify parasite species infecting human and animal populations in specific areas (Hassan et al., 2021; Khurana et al., 2021; Rodpai et al., 2018).

Next-generation sequencing (NGS) strategies have significantly advanced the study of microbial diversity (Hino et al., 2016a; Rusiñol et al., 2020a). These tools enhance taxonomic resolution by enabling the simultaneous detection of multiple taxonomic groups through direct DNA sequencing from samples (metagenomic) or PCR-based amplification of specific genetic markers (metataxonomic). Metataxonomic approaches have become invaluable in microbial ecology due to their cost-effectiveness and capacity to identify a broad range of organisms within a single sample. These studies typically target ribosomal RNA gene regions, such as 16S for bacteria, 18S for eukaryotes, and the internal transcribed spacer (ITS) region (Breitwieser et al., 2019). While widely applied in bacterial research, the use of metataxonomic in parasitology remains limited. Applications in protists have shown promising results (Garcia-Montoya et al., 2023a; Hino et al., 2016b; Moreno et al., 2018a; Rusiñol et al., 2020b; Stensvold et al., 2020; Zahedi et al., 2019), yet its implementation for human associated helminth detection is still scarce (Lappan et al., 2019).

Acknowledging the persistent limitations of conventional methods for diagnosing intestinal parasitic infections—particularly in terms of sensitivity and species-level resolution—this study explores the application of a metataxonomic approach coupled with phylogenetic methods for the simultaneous detection and, when possible, species-level confirmation of multiple parasite taxa in human fecal samples from Colombia. By evaluating a scalable, high-throughput strategy, the study aims to contribute to ongoing surveillance efforts and support more accurate monitoring in endemic settings.

## MATERIAL AND METHODS

### Sample description and microscopic diagnosis

A total of 65 fecal samples were selected in the present study based on confirmed microscopic diagnoses of the soil transmitted helminths and protists: *Ascaris*, *Trichuris*, Hookworm, *Strongyloides stercoralis, Entamoeba spp, Giardia*, *Blastocystis*, and *Endolimax*. Fecal samples were obtained from individuals in selected regions of Colombia as part of previous research projects. At the time of collection, stool samples were examined by direct microscopy following a concentration step using the MiniParasep® SF fecal parasite concentrator (Apacor Ltd, England). Samples were stored at −80 °C until DNA extraction. Prior to molecular analysis, all samples were anonymized and coded to ensure the confidentiality of participants. A complete description of the data collected, and the parasitological results is presented in Supplementary Table 1.

### DNA extraction and metataxonomic experiment

DNA extraction was performed with the Stool DNA isolation kit NORGEN BIOTEK CORP. DNA quantitation and UV spectral analysis was performed with Nanodrop 2000c to assess DNA purity and quantity.Thequantity. The V4 region of the eukaryotic rDNA gene was selected as marker for this study using the primer pair: 18S-V4Fw: CCAGCAGCCGCGGTAATTCC (Choi & Park, 2020), and the reverse primer was 18S–V4 Rev.: RCYTTCGYYCTTGATTRA, As described elsewhere (Rozo-Montoya, Bedoya-Urrego, & Alzate, 2023)

### Bioinformatic processing

Amplicon libraries were prepared and sequenced at Macrogen (Seoul, Korea) in a MiSeq (Illumina) platform to generate paired end reads of 150 bases. Amplicon reads underwent processing using the MOTHUR pipeline version 1.44 (Schloss et al., 2009). Briefly, Paired-endPaired end (PE) reads were merged using Mothur’s command “make.contigs”. Sequences with homopolymers longer than 8, with ambiguous bases, or sequences shorter than 250 bases were filtered out. The chimeric sequences were detected with VSEARCH (Rognes et al., 2016) and removed from the analysis. Read clustering to operational taxonomic units (mOTUs) was performed with the subroutine “dist.seqs” at 97% nucleotide identity. Library size for each sample was normalized with the “totalgroup” method.

The taxonomic assignment of the generated mOTUs was conducted by comparisons with the SILVA v138 ribosomal database (Quast et al., 2012) in the MOTHUR pipeline. mOTUs identified as protist or nematode were retained for subsequent alignment with the BLASTN tool (Altschul et al., 1990). We selected good quality sequences with a length ≥1000 bp and considered valid candidate mOTUs for subsequent phylogenetic analysis those that showed a bit score ≥400 in the BLASTN.

To perform the taxonomic assignation, a database was built with mOTUS harbouring sequences of the of interest present in the stool samples and a curated reference sequence database of 18S rDNA genes of the nematode and protists. The reference protist included species of the genus *Entamoeba*, *Endolimax*, *Iodamoeba*, *Cystoisospora, Besnoitia, Hyaloklossia, Caryospora, Eumonospora, Hammondia, Neospora, Toxoplasma, Nephroisospora* and *Blastocystis* subtypes (ST1 to ST17). We selected nematodes in the superfamily Ancylostomatidae (*Rhabditella axei*, *Angiostrongylus cantonensis*, *Angiostrongylus costaricensis*, *Angiostrongylus vasorum*, *Haemonchus contortus*, *Ancylostoma duodenale* and *Necator americanus)*, Dorylaimia subclass (*Soboliphyme baturini*, *Trichuris muris*, *Trichuris suis*, *Trichuris trichiura*, *Trichinella spiralis*, *Trichinella pseudospiralis)*, Strongyloididae family (*Strongyloides papillosus*, *Strongyloides ransomi*, *Strongyloides ratti* and *Strongyloides stercoralis)*, Spirurina (*Toxocara vitulorum*, *Toxocara canis*, *Toxocara cati*, *Ascaris lumbricoides*, and *Ascaris suum)*. The accession number for the reference sequences can be found in Supplementary Table 2.

The reference sequences of each taxonomic group, along with the mOTUs identified by BLASTN comparisons as candidate helminth and protists species were aligned with MAFFT v7.215 (Katoh & Standley, 2013) software. The phylogenetic tree was calculated using the IQ-TREE2 software (Minh et al., 2020) with 1000 ultrafast bootstrap pesudoreplicates to test the topology of the trees (Hoang et al., 2018). The ModelFinder was used to automatically select the best substitution model for each taxa dataset. (Kalyaanamoorthy et al., 2017).

After the confirmation of the taxonomic assignment for each protist and nematode mOTU, a database was elaborated to summarize the findings from both the microscopic smear analysis and the NGS metataxonomic analysis. The presence of a classified mOTU in any individual was noted as a qualitative value of “present.” Individuals in which the nematode mOTU was absent were categorized as “negative.” Bar graphs were generated to present the observed results in R with the GGPLOT2 package.

### Ethical considerations

Fecal samples were collected in two time periods: April-November 2015 and September 2022-February 2023 as a part of independent research projects. The use of archived, anonymized fecal samples in this study was approved by the Bioethics Committee of Sede Investigación Universitaria-Universidad de Antioquia (approval code CBE SIU 250, Date: October 18, 2023), in accordance with institutional and national guidelines. The authors had access to epidemiological data from the population; however, all information was anonymized, and participants provided written informed consent prior to data collection. No new samples were collected, and informed consent was not required due to the retrospective and anonymized nature of the material.

### Raw data availability

Raw read sequences generated in this study have been deposited in the NCBI SRA database under BioProject accession number PRJNA1293464. Reviewers can access the data using the following link: https://dataview.ncbi.nlm.nih.gov/object/PRJNA1293464?reviewer=5a293klh5f2p0atf35mq k6j2a8

## Results

### Parasitic Diagnosis Using Next-Generation Sequencing and Direct Examination

This study aimed to compare the effectiveness and performance of parasite detection in human fecal samples using two approaches: traditional smear microscopy performed by trained parasitologists, and a metataxonomic method targeting the V4 hypervariable region of the eukaryotic 18S rDNA. Previous tests confirmed that the primers used in this study are effective for detecting protists and nematodes but do not amplify DNA from cestodes and trematodes. As a result, these groups were excluded from the scope of our analysis. Accordingly, our study focused on the detection of human intestinal protists and nematodes.

The study population was selected due to the high prevalence of intestinal parasitic infections in the region, particularly geohelminths. This selection enabled a robust comparison between the performance of traditional microscopy and metataxonomic (NGS-based) approaches. We began by focusing on nematode infections. Three samples that tested negative for nematodes by microscopy were included as controls. Notably, one of these was negative for all parasites by both methods—NGS and microscopy—serving as a negative reference.

Based on microscopy results, 95.4% of individuals (62 out of 65) were positive for at least one nematode species. In comparison, the metataxonomic approach using NGS detected nematodes in 86.2% of individuals (56 out of 65). As expected, the geohelminths *Trichuris*, *Ascaris*, and hookworms were the most prevalent parasites detected. Notably, hookworms surpassed *Ascaris* in prevalence, displacing it from its typical second position in Colombia. Interestingly, *Strongyloides* was also detected, despite being less commonly reported. When comparing the performance of the two detection methods, microscopy outperformed the metataxonomic NGS approach in identifying *Ascaris* by one additional positive sample and *Trichuris* by twelve. In contrast, both methods yielded the same results for hookworm detection. Strikingly, NGS substantially outperformed microscopy in the detection of *Strongyloides*, identifying three positive samples compared to just one by microscopy— representing a 300% increase (Figure 1).

**Figure 1.**
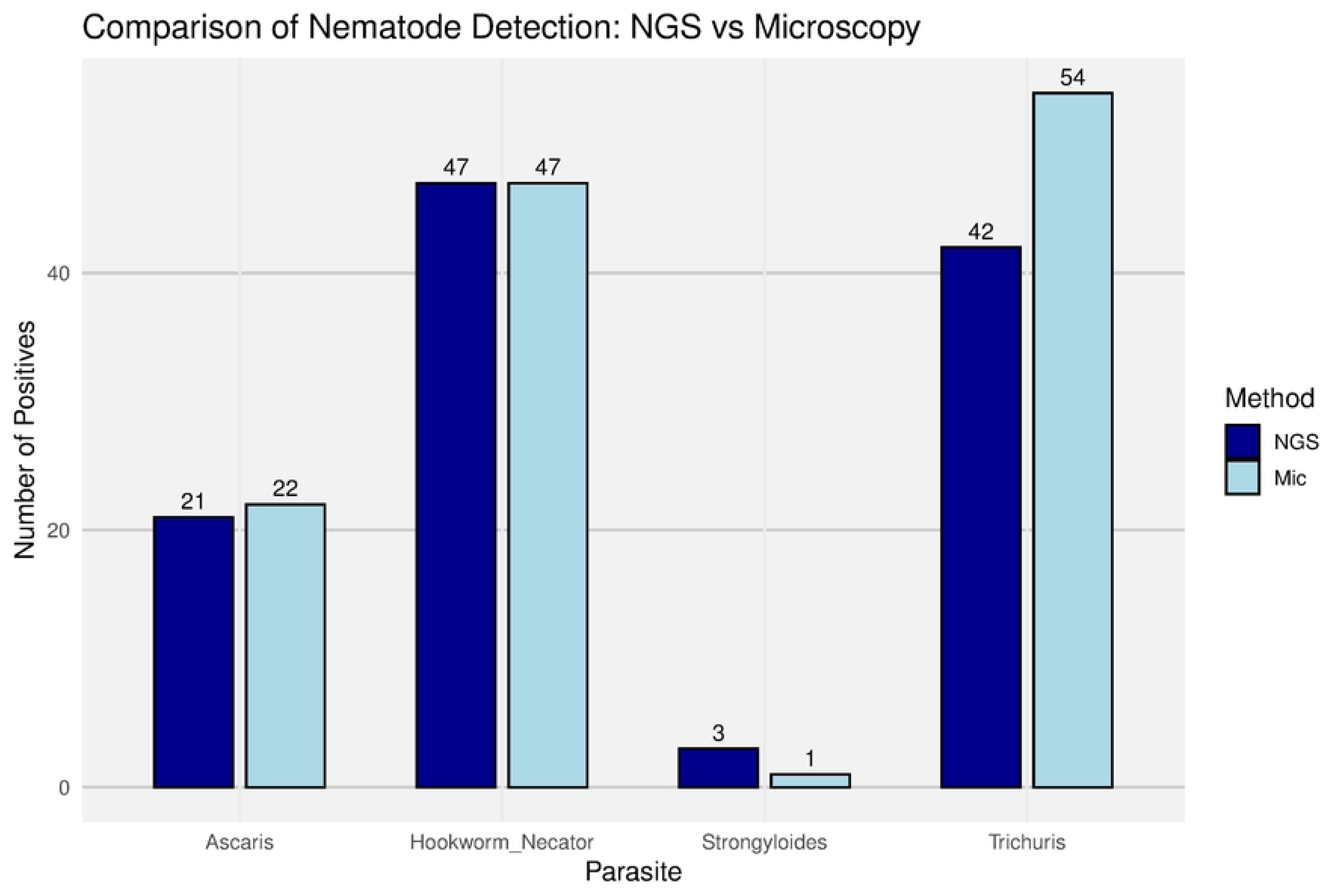
Comparison of nematode detection frequencies between NGS-Metataxonomics and microscopy-based diagnostics. Bar plots show the number of positive samples for four relevant geohelminths—*Ascaris*, *Necator*, *Strongyloides*, and *Trichuris*—detected using either molecular (NGS) or traditional microscopy approaches (Mic). Detection was binarized (positive vs. negative) and frequencies (Y-axis) were calculated across all samples in the dataset. Bars are grouped by parasite and color-coded by detection method (dark blue for NGS and light blue for microscopy).

### Phylogenetic species analysis of nematodes

To improve the taxonomic resolution of nematode identification, we performed phylogenetic analyses using the molecular operational taxonomic units (mOTU) sequences. Curated reference sequences for each of the four detected nematode genera were included to ensure accurate and reliable taxonomic assignment.

*Trichuris* mOTUs were confirmed as *T. trichiura* based on phylogenetic analysis, as they formed a well-supported monophyletic clade with curated reference sequences of *T. trichiura*, receiving a UFB (ultrafast bootstrap) support value of 96 (Figure 2).

**Figure 2.**
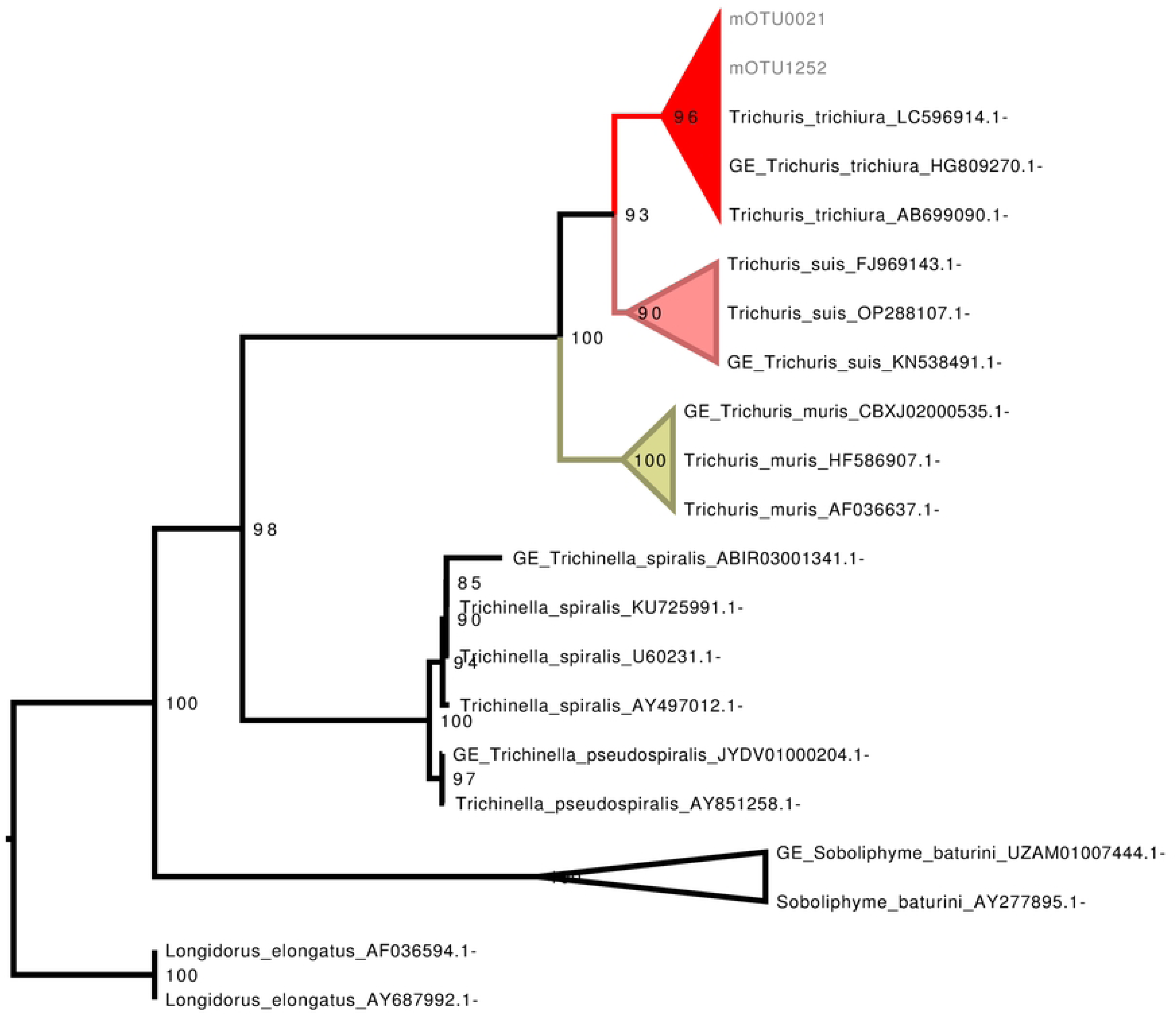
Phylogenetic analysis of *Trichuris* mOTUs for taxonomic assignment. Maximum-likelihood phylogenetic tree based on the SSU rRNA gene, constructed using a curated selection of Dorylaimida nematode sequences, including representatives of the genus *Trichuris*. The tree was inferred with 1,000 ultrafast bootstrap (UFBoot) replicates to assess branch support. Molecular OTUs (mOTUs) identified in this study are labeled with the prefix “mOTU”. Ultrafast bootstrap values are shown at the corresponding nodes. *Longiorus elongatus* was used as the outgroup.

The phylogenetic analysis of mOTUs assigned to hookworms successfully identified them as *Necator americanus*. The resulting phylogenetic tree showed a well-supported clade containing the human-infecting genera *Ancylostoma* and *Necator*. Reference sequences of *Ancylostoma* formed a distinct branch with 100% ultrafast bootstrap (UFB) support; however, none of the mOTUs from our samples clustered within this branch. Instead, all mOTUs grouped with *N. americanus* reference sequences, forming a clade with 87% UFB support. Based on this phylogenetic placement, we assigned the detected mOTUs to *N. americanus*. In conclusion, all individuals in this study who tested positive for hookworms were infected only with *Necator americanus*. (Supplementary Figure 1). Similarly, *Ascaris* mOTUs were confirmed to belong to the genus *Ascaris*, forming a clade with an ultrafast bootstrap (UFB) support value of 92. However, species-level assignment was not possible, as the 18S rDNA sequences of the closely related species *A. lumbricoides* and *A. suum* are highly similar, making it impossible to resolve them using this molecular marker (Figure 3).

**Figure 3.**
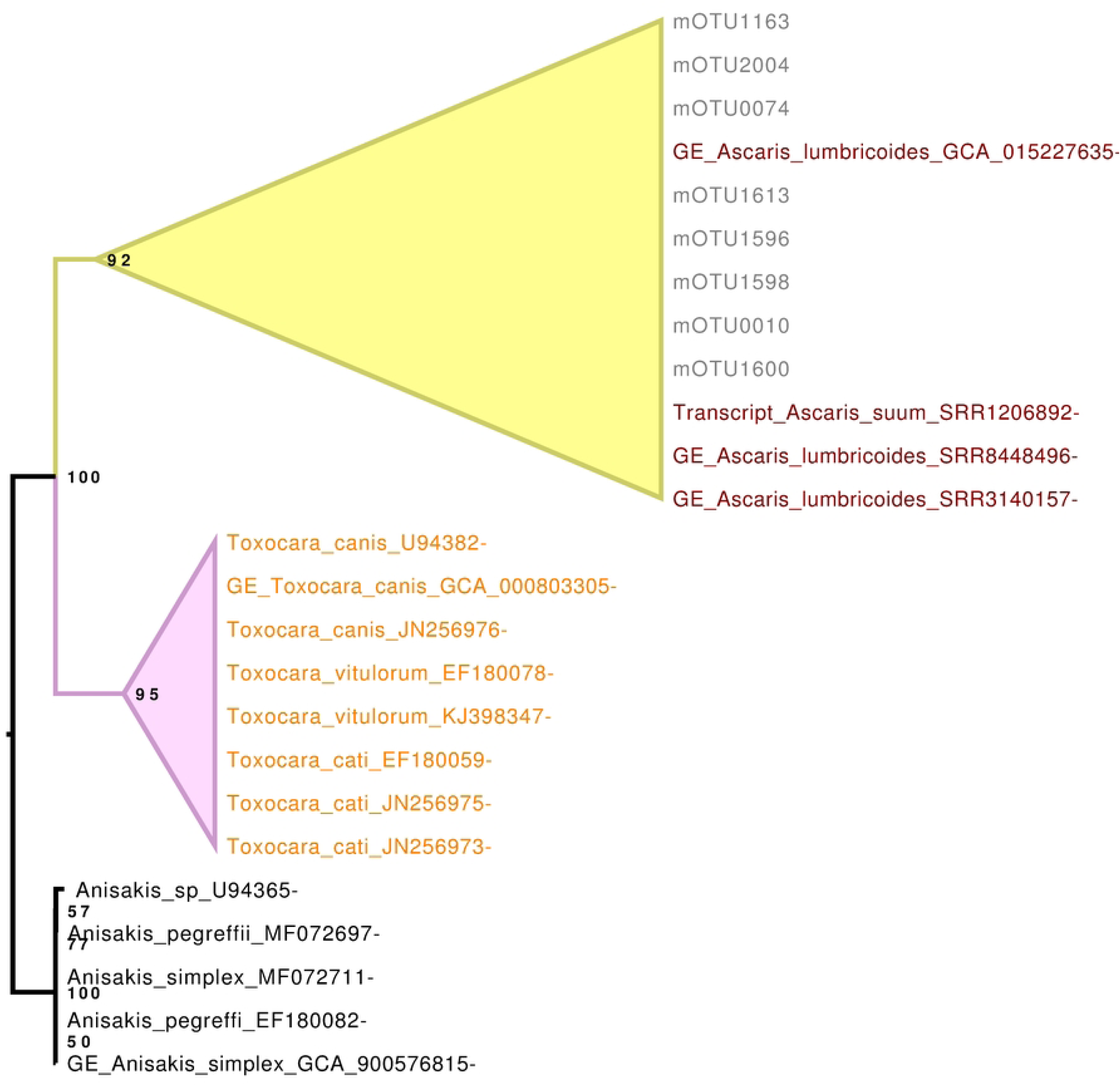
Phylogenetic analysis of Ascaris mOTUs for taxonomic assignment. Maximum-likelihood phylogenetic tree based on the SSU rRNA gene, constructed using a curated selection of nematode sequences from the genera *Ascaris*, *Toxocara* and *Anisakis*. The tree was inferred with 1,000 ultrafast bootstrap (UFBoot) replicates to assess branch support. Molecular OTUs (mOTUs) identified in this study are labeled with the prefix “mOTU”. Ultrafast bootstrap support values are shown at the respective nodes. *Anisakis* genus was used as the outgroup.

Finally, the *Strongyloides* sequences from the three infected individuals clustered into a single mOTU, which was confidently assigned to *S. stercoralis*. This mOTU formed a well-supported clade with 100% ultrafast bootstrap (UFB) support alongside curated reference sequences of *S. stercoralis*, confirming its taxonomic identity. Interestingly, one sample was positive for *S. stercoralis* by microscopy, based on the visualization of larvae, but was classified as negative for this species by NGS. Instead, NGS identified this sample as positive for *Necator americanus*. Remarkably, all three *S. stercoralis* infections were exclusively detected by NGS, highlighting its enhanced sensitivity for detecting this parasite (Figure 4).

**Figure 4.**
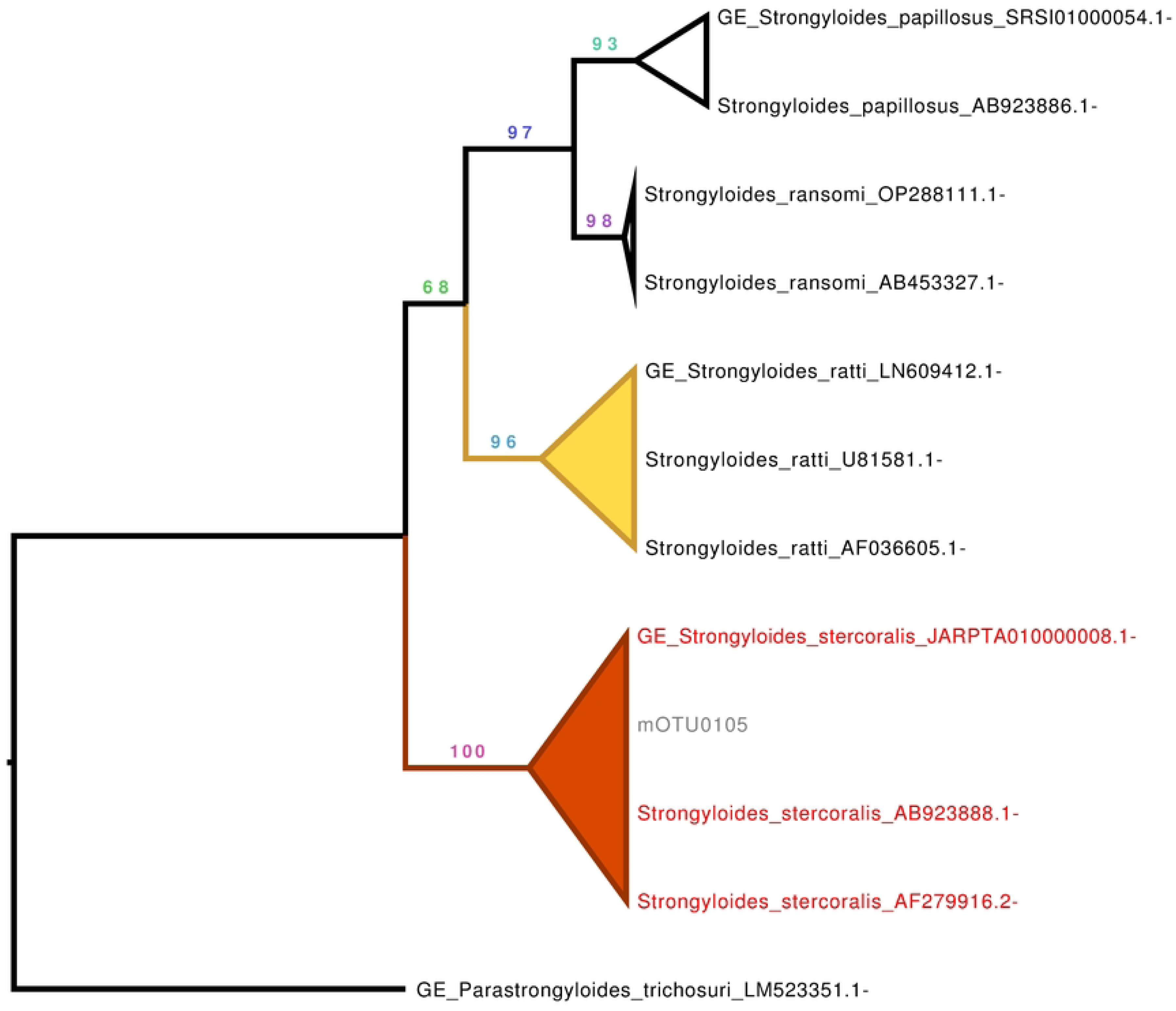
Phylogenetic analysis of *Strongyloides* mOTUs for taxonomic assignment. Maximum-likelihood phylogenetic tree based on the SSU rRNA gene, constructed using a curated selection of *Strongyloides* species sequences. The tree was inferred with 1,000 ultrafast bootstrap (UFBoot) replicates to assess branch support. Molecular OTUs (mOTUs) identified in this study are labeled with the prefix “mOTU”. Ultrafast bootstrap support values are shown at the corresponding nodes. *Parastrongyloides trichosuri* was used as the outgroup.

Based solely on the NGS results—given their higher specificity—we found that 32.1% of individuals harbored single nematode infections, while 67.9% presented with mixed infections. The most common coinfection was *N. americanus* + *T. trichiura* (n = 22), followed by *N. americanus* + *T. trichiura* + *Ascaris* (n = 12). Remarkably, one individual was infected with all four detected nematode species. Notably, no single infections with *S. stercoralis* were observed; this species was detected only in individuals co-infected with other geohelminths (Figure 5).

**Figure 5.**
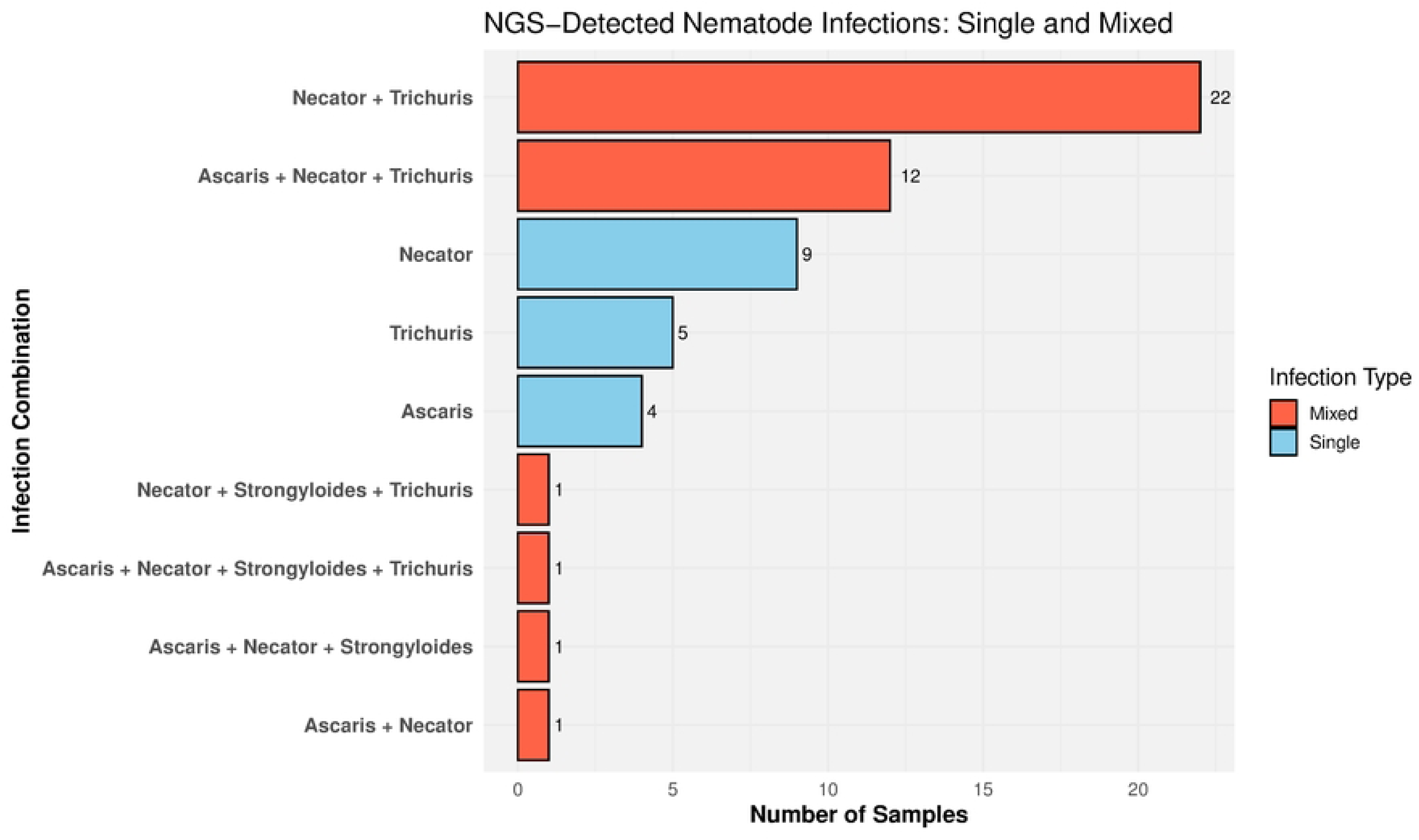
Distribution of single and mixed nematode infections detected by Next-Generation Sequencing (NGS). Horizontal bar plot summarizing infection combinations detected by NGS across all samples for four soil-transmitted helminths: *Ascaris spp.*, *Necator americanus*, *Strongyloides stercoralis*, and *Trichuris trichuria*. Each bar represents the number of samples exhibiting a specific infection pattern (X-axis), categorized as either single-species (Single) or mixed-species infections (Mixed). Bars are color-coded by infection type: blue for single infections and red for mixed infections.

### *Entamoeba* and Other Amoebae

In the NGS metataxonomic analysis, species-level assignment was performed using a phylogenetic approach. This allowed us to confidently confirm the presence of *E. coli*, *E. hartmanni*, and *E. dispar* in the studied population, with strong phylogenetic support (UFB ≥ 98) for each clade (Supplementary Figure 2).

A total of 34 individuals (52.3%) tested positive for any *Entamoeba* species by microscopy, while 42 individuals (64.6%) were positive using NGS-based metataxonomics. Overall, NGS outperformed microscopy in detecting *Entamoeba* species. The most striking difference was observed for *E. hartmanni*, with only 9 positives identified by microscopy compared to 29 detected by NGS—over three times more. For *E. coli*, the results were more similar, with NGS detecting 25 positives and microscopy 24. In the case of *E. histolytica/dispar/moshkovskii*, both methods yielded identical results, with 13 positive individuals each (Figure 6). Interestingly, the phylogenetic analyses resolved the amoeba diversity at the species level, with high bootstrap support distinguishing closely related taxa, including members of the Entamoeba histolytica/dispar/moshkovskii complex.

**Figure 6.**
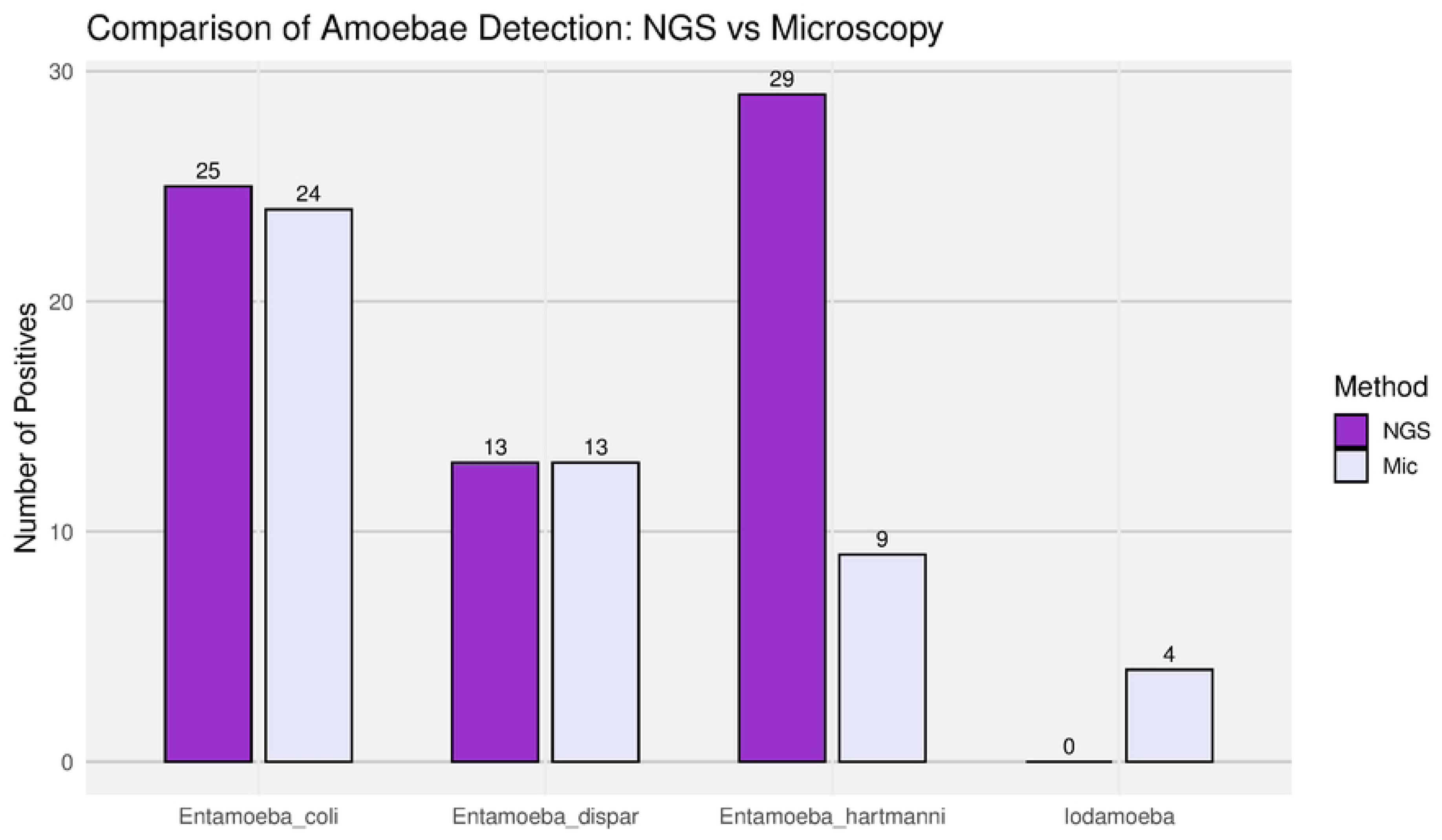
Comparison of amoebae detection using NGS-Metataxonomics and microscopy. Bar plot showing the number of positive samples for intestinal amoebae: *Entamoeba dispar*, *Entamoeba coli*, *Entamoeba hartmanni*, and *Iodamoeba* sp., detected using either NGS-Metataxonomics (NGS) or microscopy (Mic). Bars represent the total number of detections for each parasite by method across all samples (Y-axis).

The microscopy examination identified four samples positive for *Iodamoeba*, whereas no detections were observed using the NGS-based approachmicroscopy identified 4 positive samples for *Iodamoeba*, whereas NGS detected none. This discrepancy is likely due to poor primer binding, as the reverse primer used for PCR shows limited annealing efficiency in *Iodamoeba* due to low sequence conservation in the targeted rDNA region.

Based on the higher specificity of the NGS results, we examined the distribution of *Entamoeba* species and their co-occurrence patterns within the studied population. While single-species infections involving any of the three detected *Entamoeba* species (*E. coli*, *E. hartmanni*, and *E. dispar*) were observed, mixed infections involving all possible species combinations were also detected. The most common infection types were single *E. hartmanni* infections and mixed *E. coli* + *E. hartmanni* infections, with 12 cases each (Figure 7).

**Figure 7.**
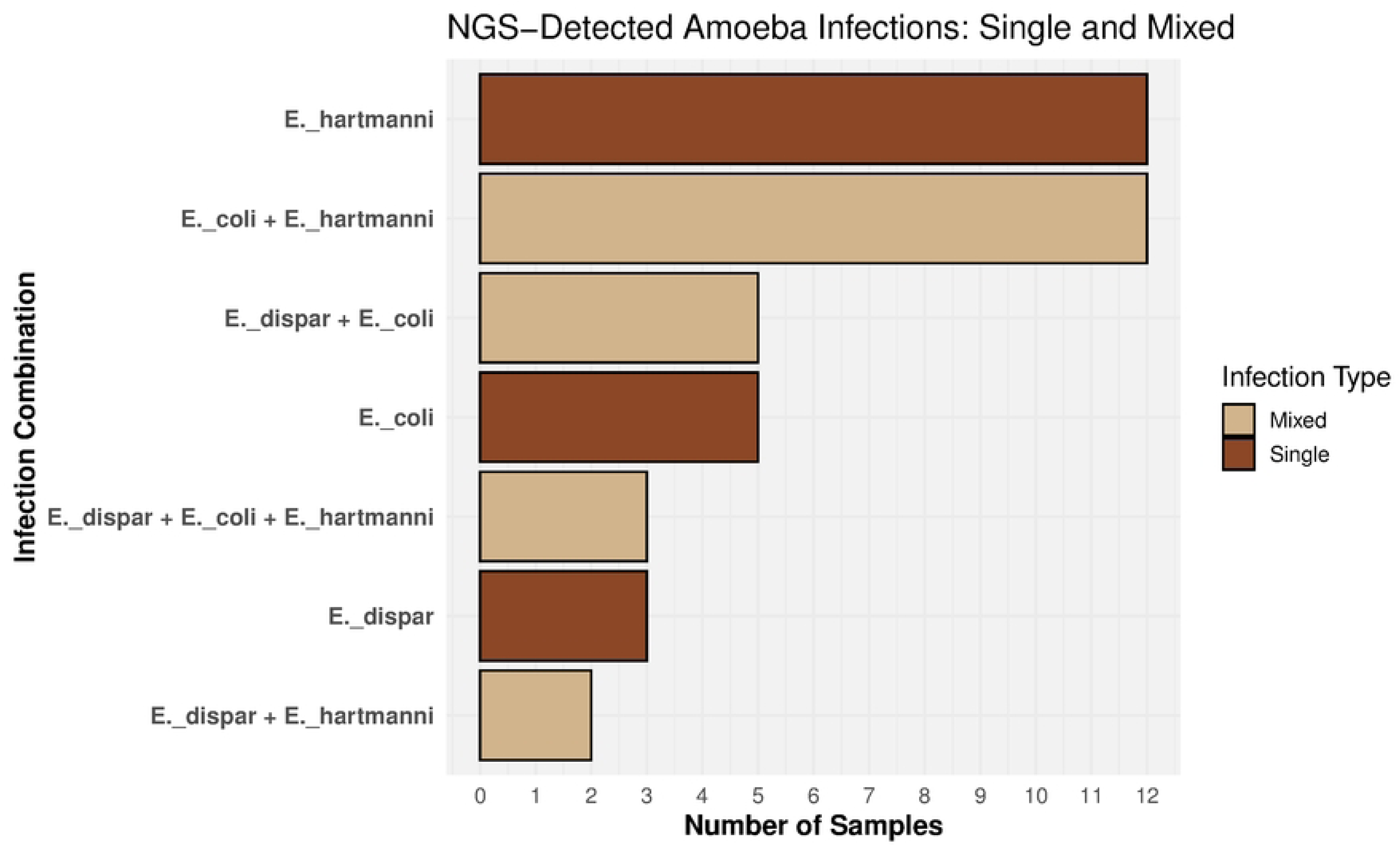
Distribution of single and mixed amoeba infections detected by NGS-Metataxonomics. Horizontal bar plot summarizing infection combinations detected by NGS across all samples for intestinal amoebae: *Entamoeba dispar*, *Ent. coli*, *Ent. hartmanni*. Each bar represents the number of samples exhibiting a specific infection pattern (X-axis), categorized as either single-species (Single) or mixed-species (Mixed) infections. Bars are color-coded by infection type: brown for single infections and tan for mixed infections. Parasite names were abbreviated for clarity (e.g., Entamoeba → E.).

#### Blastocystis

*Blastocystis* represents another compelling case in which microscopy significantly underestimated the number of colonized individuals. While Parasep concentration and direct stool smear analyses detected only 4 positive samples, the NGS metataxonomic approach identified *Blastocystis* in 46 individuals (70.7%), an eleven-fold increase (Figure: Bars_Blastocystis) (Figure 8).

**Figure 8.**
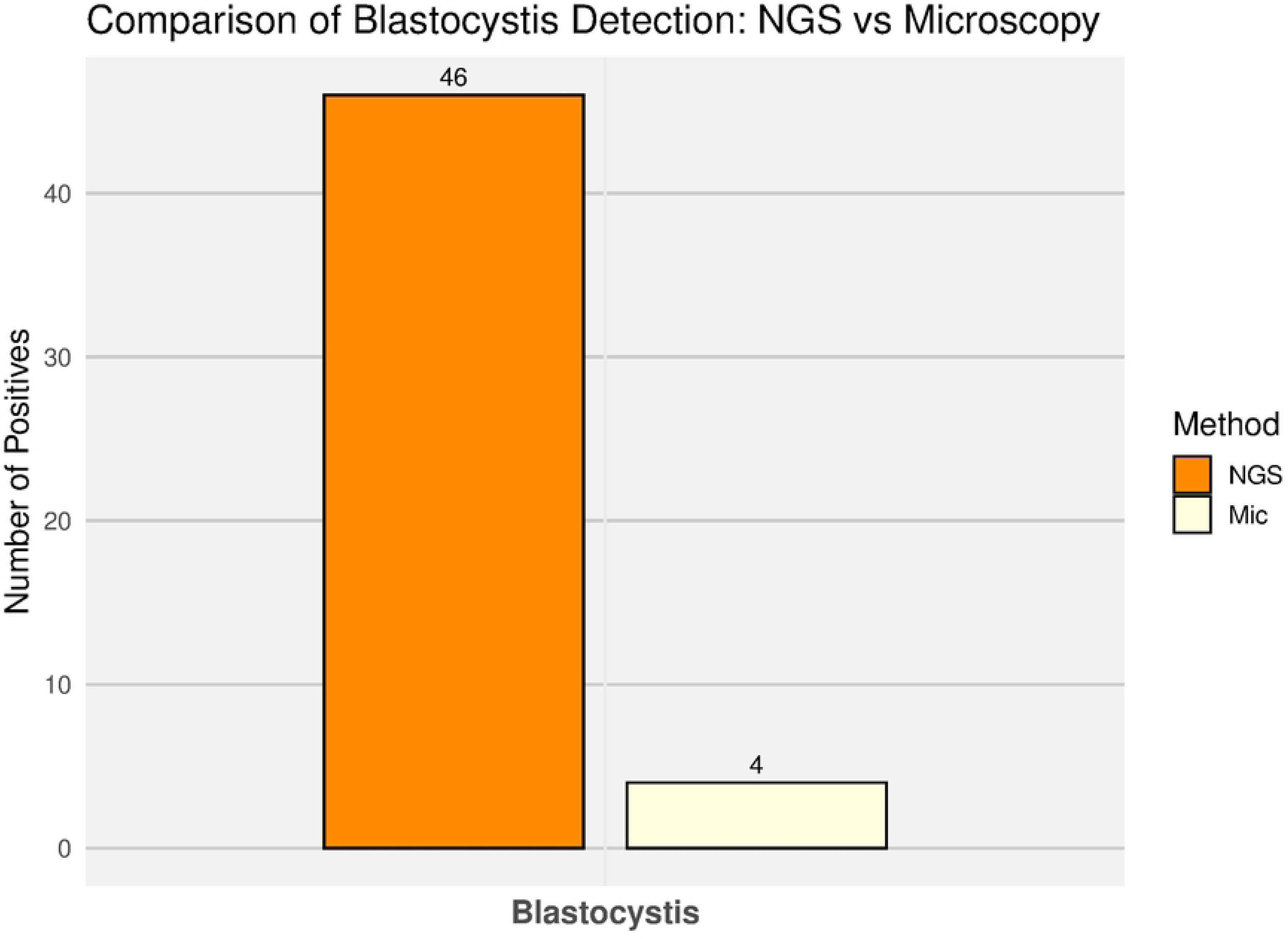
Comparison of *Blastocystis* detection using NGS-Metataxonomics and Microscopy. Bar plot showing the number of samples positive for *Blastocystis hominis* detected by either NGS-Metataxonomics (NGS) and by conventional microscopy (Mic). Each bar represents the total number of positive samples identified by each diagnostic method (Y-axis).

Subtype classification was performed as described in the Methods section, using the mOTUs assigned to *Blastocystis*. As shown in the phylogenetic tree (Supplementary Figure: Blastocystis_tree), all detected mOTUs were confidently assigned to subtypes ST1, ST2, and ST3, with robust phylogenetic support (UFB 100) (Supplementary Figure 3).

The most frequent colonization pattern involved a single subtype: ST1 (n = 13), followed by ST3 (n = 10) and ST2 (n = 9). Mixed subtype colonization was observed in 14 individuals (21.5%), with ST1+ST3 being the most common combination (n = 8). Notably, triple subtype colonization (ST1+ST2+ST3) was detected in 2 individuals (Figure 9).

**Figure 9.**
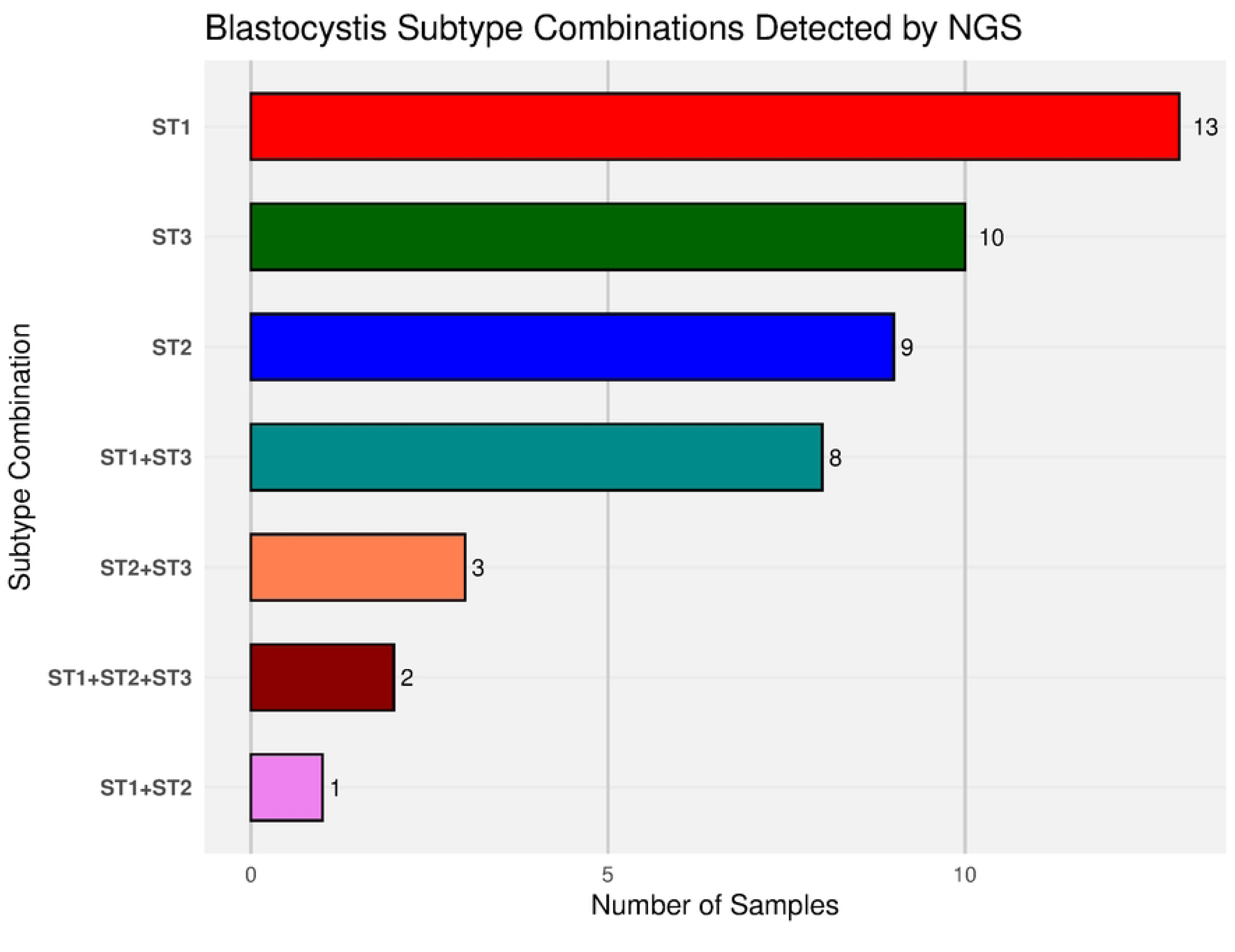
Distribution of Blastocystis subtype (ST) combinations detected by NGS-Metataxonomics. The bars represent the occurrence of single and mixed Blastocystis subtypes among positive samples (X-axis).

### Other Protozoa

Regarding the other protozoan intestinal parasites, the NGS metataxonomic approach detected only *Cystoisospora*. Four mOTUs were assigned to this coccidian genus. Phylogenetic analysis showed that these mOTUs formed a monophyletic group with known mammalian parasitic species including *C. belli*, *C. ohionensis*, *C. suis*, *C. canis*, and *C. felis*, with strong bootstrap support (98%) (Figure 10). This phylogenetic placement confidently confirmed the genus-level identity; however, resolution at the species level was not possible based on the 18S rDNA marker used. Microscopy reported the presence of *Chilomastix*, *Cyclospora*, and *Giardia*, while NGS metataxonomics detected only *Cystoisospora*, which was found in two individuals. Interestingly, the two *Cystoisospora*-positive individuals were negative for *Cyclospora*, the other coccidian parasite (Figure 11).

**Figure 10.**
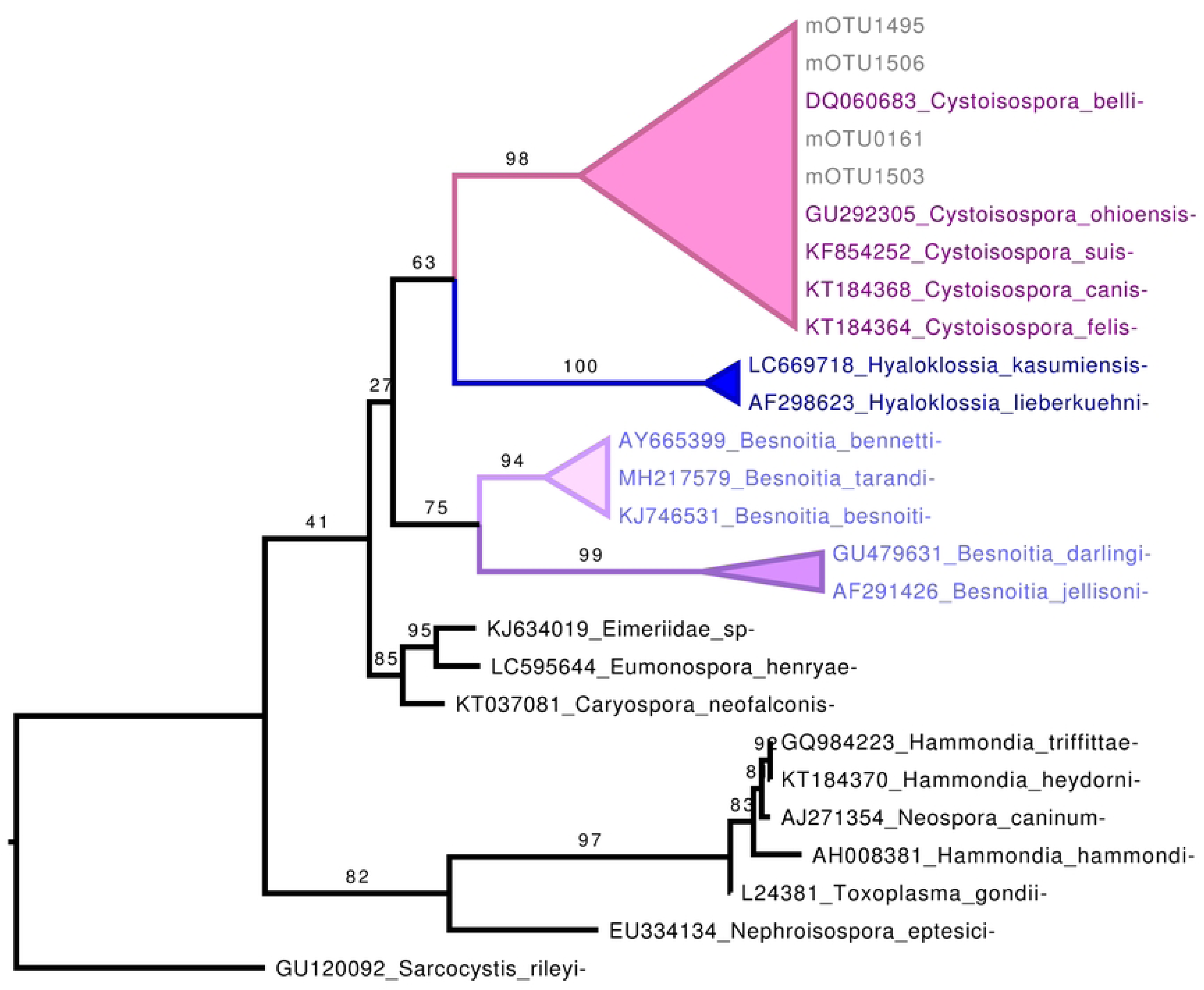
Phylogenetic analysis of *Cystoisospora* mOTUs for taxonomic assignment. Maximum-likelihood phylogenetic tree based on the 18S rRNA gene, constructed using a curated selection of reference sequences from apicomplexan parasites, including representatives of the genus *Cystoisospora*. The tree was inferred with 1,000 ultrafast bootstrap (UFBoot) replicates to assess branch support. Molecular OTUs (mOTUs) identified in this study are labeled with the prefix “mOTU”. Ultrafast bootstrap support values are shown at the corresponding nodes. *Sarcocystis rileyi* was used as the outgroup.

**Figure 11.**
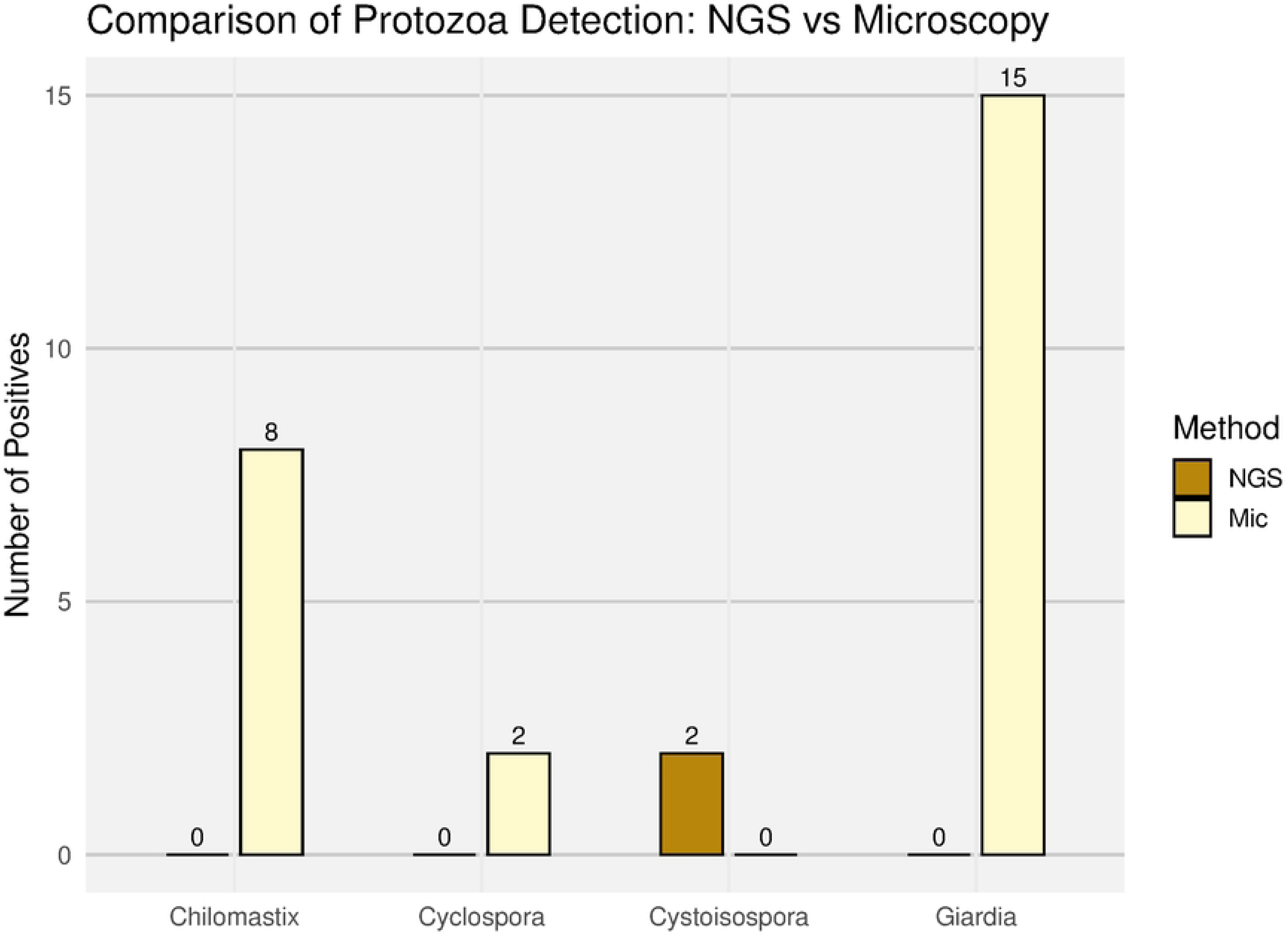
Comparison of other protozoan parasite detection by NGS-Metataxonomics and microscopic smear analysis. Bar plot summarizing the number of positive samples for other intestinal protozoa (*Giardia*, *Cyclospora*, *Cystoisospora*, and *Chilomastix*) as detected by NGS-Metataxonomics (NGS) and microscopy (Mic). Each bar represents the total number of samples testing positive per parasite and method (Y-axis).

## Discussion

Intestinal parasites remain a significant public health burden in low- and middle-income countries and are increasingly recognized as an emerging concern in developed nations (Fletcher et al., 2012; Li et al., 2020; Lucas Dato et al., 2024a). Since the initial discovery of intestinal protists and nematodes, microscopy has served as the foundation for the diagnosis of intestinal parasitic infections in both humans and animals (Soares et al., 2020; van Lieshout & Roestenberg, 2015) and due to the challenges associated with culturing many intestinal parasites under controlled laboratory conditions (Feix et al., 2023), still persist in many laboratories as the primary method for taxonomic classification.

The advent of PCR in the 1980s and 1990s marked a major advancement, improving the sensitivity and specificity of diagnostic tests and enabling more refined classification of parasitic organisms (Baltrušis & Höglund, 2023; Garg et al., 2024; Sow et al., 2017). However, recent biotechnological innovations have continued to drive progress across all areas of the life sciences. Notably, next-generation sequencing (NGS) technologies have revolutionized parasitology by enabling high-throughput detection methods such as metataxonomic analysis (Antonopoulos et al., 2024; Basirpour et al., 2025; Mthethwa et al., 2021). Unlike traditional qPCR-based methods that rely on fluorescence detection of specific amplicons, NGS allows for the sequencing of hundreds of thousands of DNA fragments simultaneously (Basirpour et al., 2025). This dramatically enhances detection sensitivity and, when coupled with modern bioinformatic tools and phylogenetic analyses, significantly improves taxonomic resolution and specificity (Rozo-Montoya, Bedoya-Urrego, & Alzate, 2023).

In this study, we combined a metataxonomic approach with classical phylogenetic methods to detect and confidently classify intestinal parasites in rural populations of Colombia, where high prevalence rates have been historically documented (Castañeda et al., 2024)(. While metataxonomic methods have been increasingly applied for the detection of protist parasites (Fitri et al., 2022; Garcia-Montoya et al., 2023b; Kang et al., 2024; Moreno et al., 2018b), our study expanded their use to include the detection of geohelminths, including *Strongyloides stercoralis*, and enabled species-level assignment whenever possible through a curated phylogenetic framework. Additionally, we aimed to compare the performance of traditional microscopy with that of NGS-based metataxonomics, highlighting the strengths and limitations of each method in accurately characterizing parasitic infections.

Although both technologies successfully detected intestinal nematodes, notable differences were observed in certain cases. For *Trichuris trichiura*, microscopy identified significantly more infected individuals than the NGS-based metataxonomic approach. A plausible explanation is the high resistance of the *Trichuris* eggshell (Wharton & Jenkins, 1978), which may hinder efficient DNA extraction and reduce the yield of amplifiable material. A similar, albeit less pronounced, pattern was observed for *Ascaris*, with microscopy detecting one additional positive case. We applied a physical and chemical method for lysis according to manufacturer instructions. Future studies should implement improved lysis protocols to enhance the disruption of tough nematode eggshells (Waindok et al., 2022; Yoshida, 1920) and increase detection sensitivity for these species. An additional consideration with *Ascaris* is that since *A. lumbricoides* and *A. suum* are closely related species (Zhou et al., 2020), the 18S rRNA gene does not provide sufficient resolution to distinguish between them, limiting species-level assignment in metataxonomic analyses.

In contrast, for hookworms and *Strongyloides stercoralis*—both of which possess less resistant external structures (Chauhan et al., 2017; Matthews, 1986)—microscopy did not outperform metataxonomics. In fact, NGS-based metataxonomic analysis proved more effective for *S. stercoralis*, detecting three times as many positive samples. This is particularly relevant given the clinical importance of *S. stercoralis*, which can cause severe, life-threatening infections, especially in immunocompromised individuals (Lucas Dato et al., 2024b). Another advantage of using NGS technologies is their ability to reduce the risk of misidentifying *Strongyloides* rhabditoid larvae as hookworm larvae. Traditional parasitological methods for its detection (e.g., direct smear, Baermann, agar plate culture) are labor-intensive, technically demanding, and have limited sensitivity—often detecting fewer than 60–70% of cases in single-sample analyses (Chan & Thaenkham, 2023). Molecular approaches, including NGS, offer significantly improved sensitivity and can effectively overcome the challenges posed by low and intermittent larval shedding (Chan & Thaenkham, 2023).

One distinct advantage of the metataxonomic approach is its ability—when combined with phylogenetic analysis—to assign most intestinal parasites to the species level within a robust and well-supported taxonomic framework. This contrasts with microscopy, which typically permits only genus-level identification, thereby limiting our understanding of the true epidemiology, burden, and transmission dynamics of parasitic infections. Using our combined NGS metataxonomic and phylogenetic strategy, we were able to confirm that the Colombian populations studied are infected, in descending order of frequency, with *Trichuris trichiura*, *Necator americanus*, *Ascaris* spp., and *Strongyloides stercoralis*.

In the case of protist detection, the NGS-based metataxonomic approach has been successfully applied in several studies involving human stool samples and wastewater. Among the protists reliably detected using this technology are *Blastocystis*, *Giardia*, *Entamoeba*, *Balantioidesdium*, *Acanthamoeba*, *Cryptosporidium*, *Dientamoeba*, and *Rhogostoma* (Garcia-Montoya et al., 2023b; Rozo-Montoya, Bedoya-Urrego, & alzate, 2023; Sarzhanov et al., 2021)(Moreno et al., 2018b). These examples highlight the broad taxonomic scope and sensitivity of metataxonomic methods for detecting a wide range of eukaryotic microorganisms in complex biological and environmental samples. As shown in previous studies worldwide (Fusaro et al., 2024; Garcia-Montoya et al., 2023b; Jiménez et al., 2019; Osorio-Pulgarin et al., 2021; Tito et al., 2019), *Blastocystis* is the most prevalent intestinal protist in humans. Our study reflected the same trend, with *Blastocystis* detected in nearly 71% of the tested population using NGS-based metataxonomic technologies. Strikingly, the sensitivity of microscopic methods was notably lower, as metataxonomics detected over eleven times more colonized individuals than traditional smear techniques.

Another clear advantage of the Metataxonomics+Phylogenetic approach is the ability to assign *Blastocystis* to subtypes and to detect mixed-subtype infections (Garcia-Montoya et al., 2023b). Our findings align with previous studies by our group and others, indicating that the dominant subtypes colonizing Colombian populations are ST1, ST2, and ST3, with ST1 being the most prevalent. Additionally, we found that mixed-subtype colonization was common, occurring in approximately 22% of positive individuals. The most frequent combination was ST1+ST3, and two individuals harbored all three subtypes simultaneously.

The second most prevalent genus detected was *Entamoeba*, with nearly 65% of individuals testing positive by NGS-based metataxonomics. In contrast, microscopic smear analysis detected *Entamoeba* in 52% of the individuals. The most striking discrepancy between the two methods was observed at the species level: in our study, *E. hartmanni* appeared to be underdiagnosed by microscopy, with NGS detecting three times more cases. This underestimation may be due to morphological misidentification or confusion with other protists under light microscopy (Servián et al., 2024). For *Entamoeba coli* and *E. dispar*, both methods showed similar performance. In microscopy, the amoeba cysts identified as part of the *Entamoeba histolytica/dispar/moshkovskii* complex were classified as *Entamoeba dispar* in the NGS analysis, thanks to the resolution provided by phylogenetic inference. The similar number of positive samples obtained by both methods suggests comparable performance in detecting this genus.

An additional discordance was noted with *Iodamoeba*: microscopy reported four positive cases, whereas NGS-based metataxonomics failed to detect any. We suspect this is due to inefficient amplification caused by mismatches between the reverse primer and the 18S-V4 target region in *Iodamoeba*, leading to poor PCR performance during library preparation. Overall, the population studied showed colonization primarily by non-pathogenic, commensal amoebae. Phylogenetic analysis revealed strong resolution within the *E. histolytica/E. dispar/E. moshkovskii* species complex, and we can confidently rule out the presence of the pathogenic *E. histolytica* in the detected *Entamoeba* mOTUs. Our results agree with PCR-based molecular studies in Colombian population which have reported a higher circulation of *E. dispar* compared with *E. histolytica* (Fotedar et al., 2007; López et al., 2015).

The most incongruent results were observed for other protozoan parasites, namely *Chilomastix*, *Cyclospora*, *Cystoisospora*, and *Giardia*. For these taxa, there was no concordance between microscopy and NGS metataxonomic results; each was detected by only one method, with no mutual confirmation. In the case of *Chilomastix*, bioinformatic analysis revealed a problem similar to what we observed with *Iodamoeba*: the reverse primer target region shows poor sequence conservation in this genus, likely impairing efficient primer annealing and thus detection by PCR.

For *Cyclospora*, although we cannot entirely rule out primer incompatibility, the same primer set has successfully amplified other apicomplexan parasites (e.g., *Cystoisospora*, *Cryptosporidium*, *Sarcocystis) (Garcia-Montoya et al., 2023b)* in past analyses. Therefore, no definitive conclusion can be made regarding *Cyclospora* detection.

Interestingly, NGS did not detect *Giardia* in any of the samples analyzed. However, in previous work using the same primers and protocols, we have successfully detected *Giardia* in human stool samples from Colombia (Rozo-Montoya, Bedoya-Urrego, & Alzate, 2023)and wastewater from Colombia. This suggests that the absence of *Giardia* in the current study may reflect a true absence in the sampled population rather than a technical limitation of the method.

Other molecular approaches, such as PCR, have been employed to detect intestinal parasites. However, these techniques present inherent limitations, especially when aiming to identify a wide range of taxa. Detecting more than 20 different parasite genera—each comprising multiple species or subtypes, as in the case of *Blastocystis*—requires the use of multiple primer sets and separate reactions. This not only reduces scalability but also increases cost and complexity over time. In contrast, the NGS-based metataxonomic approach enables the simultaneous detection of a broad and potentially unlimited diversity of intestinal parasites in a single assay. Furthermore, the downstream analysis can be streamlined and automated using current, well-established bioinformatic pipelines.

Despite its advantages, our current metataxonomic approach also has known limitations. It fails to detect cestodes and trematodes, likely due to poor primer annealing caused by the high sequence divergence of their 18S rDNA V4 region compared to other intestinal parasites such as protists and nematodes. Bioinformatic analyses from this study indicate that Platyhelminths are not efficiently amplifiable with the primers used. Additionally, certain amoebae—such as *Iodamoeba*—appear to be inefficiently amplified, likely due to mismatches at the primer binding sites. These findings highlight the need for improved primer design to broaden taxonomic coverage and enhance the diagnostic utility of metataxonomic protocols.

We are now entering a new era in parasitology, empowered by an expanded molecular toolkit that includes genomic, metagenomic, and metataxonomic approaches. These technologies provide high-resolution taxonomic and evolutionary insights, enable the genomic characterization of unculturable parasites, and enhance the detection of parasitic organisms in complex samples such as stool or environmental matrices (Vatta & Cacciò, 2025).

## Data Availability

ncbi sra

## Acknowledgements

This study was funded by Escuela de Microbiología-CODI, Universidad de Antioquia, under grant code 2023-64370.

---

**Supplementary Figure 1.**
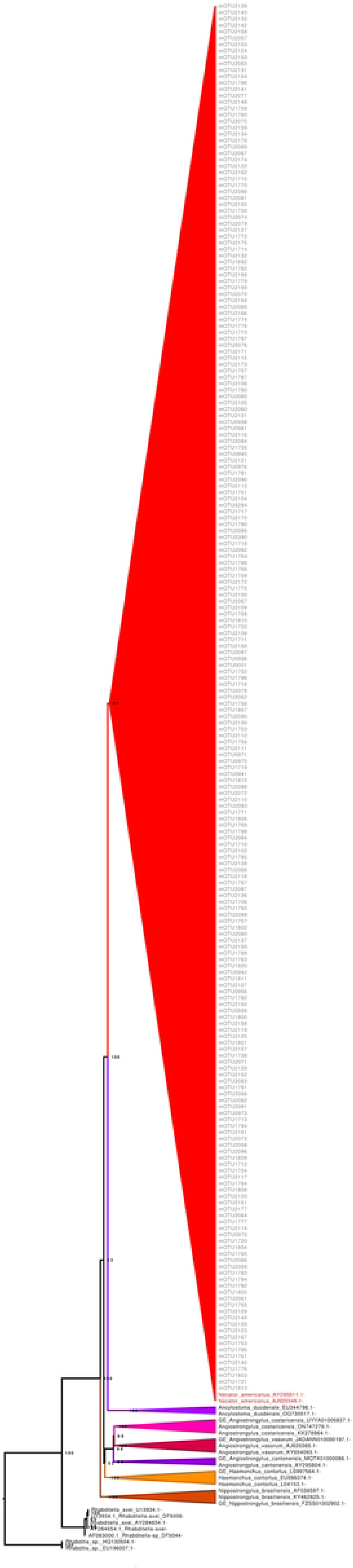
Phylogenetic analysis of *Necator* mOTUs for taxonomic assignment. Maximum-likelihood phylogenetic tree based on the 18S rRNA gene, constructed using a curated selection of reference sequences from nematode genera related to hookworms, including representatives of *Necator* and *Ancylostoma*. The tree was inferred with 1,000 ultrafast bootstrap (UFBoot) replicates to assess branch support. Molecular OTUs (mOTUs) identified in this study are labeled with the prefix “mOTU”. All hookworm mOTUs clustered within the *Necator americanus* clade, indicating that this was the sole hookworm species detected in the sampled population. Ultrafast bootstrap support values are shown at the corresponding nodes. Rhabditis sp. was used as the outgroup.

**Supplementary Figure 2.**
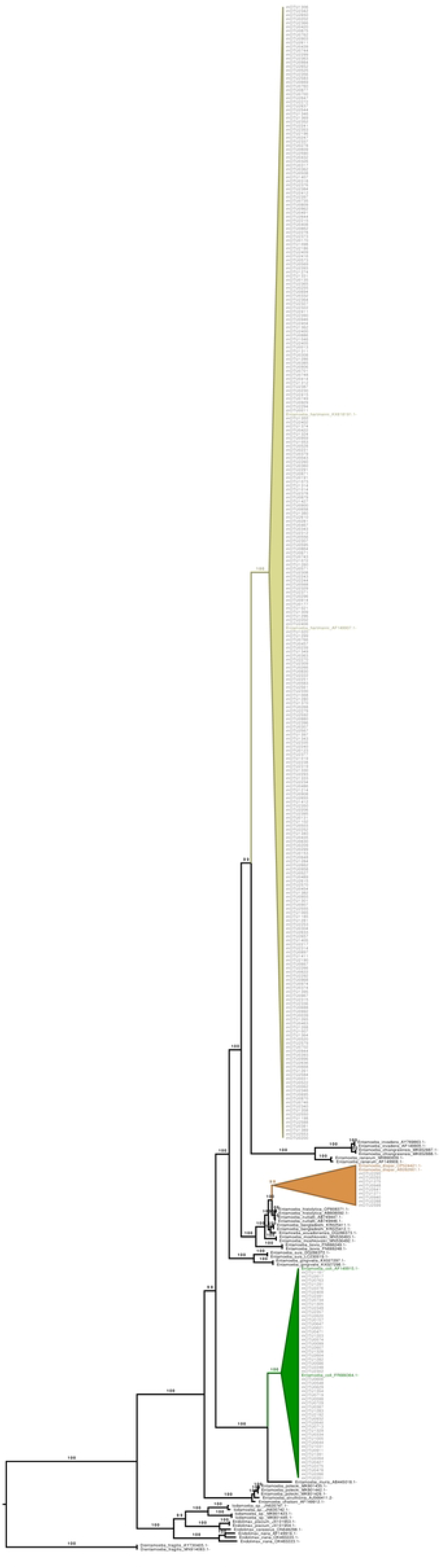
Phylogenetic analysis of *Entamoeba* mOTUs for taxonomic assignment. Maximum-likelihood phylogenetic tree based on the 18S rRNA gene, constructed using a curated selection of reference sequences from the genus *Entamoeba*. The tree was inferred with 1,000 ultrafast bootstrap (UFBoot) replicates to assess branch support. Molecular OTUs (mOTUs) identified in this study are labeled with the prefix “mOTU”. All mOTUs clustered confidently within known *Entamoeba* species clades, supporting their taxonomic assignments. Ultrafast bootstrap support values are indicated at the corresponding nodes. Representative sequences of Dientamoeba fragilis were used as the outgroup.

**Supplementary Figure 3.**
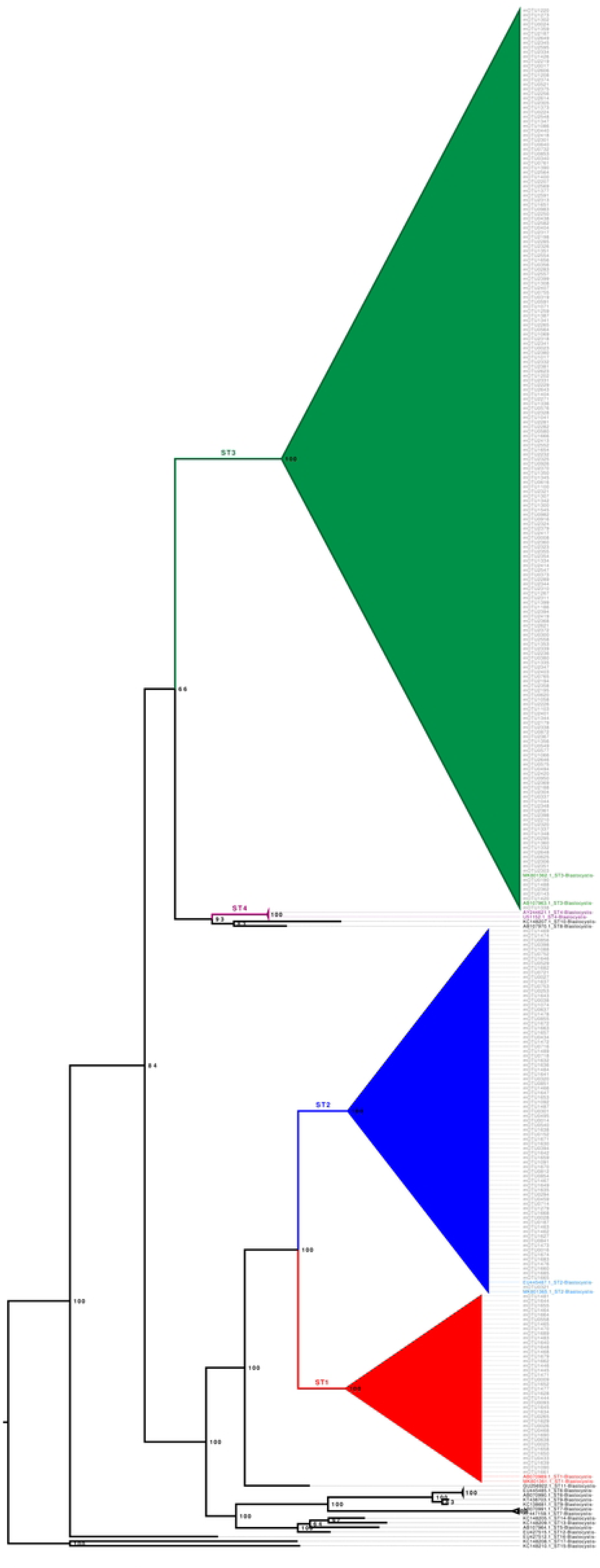
Phylogenetic analysis of *Blastocystis* mOTUs for subtype (ST) classification. Maximum-likelihood phylogenetic tree based on the 18S rRNA gene, constructed using a curated selection of *Blastocystis* reference sequences representing subtypes (STs) 1 through 17, including all subtypes commonly associated with human populations. Molecular OTUs (mOTUs) identified in this study are labeled with the prefix “mOTU”. The tree was inferred with 1,000 ultrafast bootstrap (UFBoot) replicates to assess branch support, and UFBoot values are shown at the corresponding nodes. All mOTUs clustered within well-supported clades corresponding to known *Blastocystis* subtypes, enabling confident subtype assignment. A combined clade of *Blastocystis* ST15 and ST17 was used as the outgroup.

